# Deep Learning using Susceptibility-Weighted MR Sequence to Detect Microbleeds and Classify Cerebral Small Vessel Disease

**DOI:** 10.1101/2023.03.30.23287989

**Authors:** Ruizhen Wu, Huaqing Liu, Hao Li, Lifen Chen, Lei Wei, Xuehong Huang, Xu Liu, Xuejiao Men, Xidan Li, Lanqing Han, Zhengqi Lu, Bing Qin

## Abstract

**Background:** Microbleeds (CMBs) serve as neuroimaging biomarkers to assess risk of intracerebral hemorrhage and diagnose cerebral small vessel disease (CSVD). Therefore, detecting CMBs can evaluate the risk of intracerebral hemorrhage and use its presence to support CSVD classification, both are conducive to optimizing CSVD management. This study aimed to develop and test a deep learning (DL) model based on susceptibility-weighted MR sequence (SWS) to detect CMBs and classify CSVD to assist neurologists in optimizing CSVD management.

**Methods:** Patients with arteriolosclerosis (aSVD), cerebral amyloid angiopathy (CAA), and cerebral autosomal dominant arteriopathy with subcortical infarcts and leukoencephalopathy (CADASIL) treated at three centers were enrolled between January 2017 and May 2022 in this retrospective study. The SWSs of patients from two centers were used as the development set, and the SWSs of patients from the remaining center were used as the external test set. The DL model contains a Mask R-CNN for detecting CMBs and a multi-instance learning (MIL) network for classifying CSVD. The metrics for model performance included intersection over union (IoU), Dice score, recall, confusion matrices, receiver operating characteristic curve (ROC) analysis, accuracy, precision, and F1-score.

**Results:** A total of 364 SWS were recruited, including 336 in the development set and 28 in the external test set. IoU for the model was 0.523 ± 0.319, Dice score 0.627 ± 0.296, and recall 0.706 ± 0.365 for CMBs detection in the external test set. For CSVD classification, the model achieved a weighted-average AUC of 0.908 (95% CI: 0.895-0.921), accuracy of 0.819 (95% CI: 0.768-0.870), weighted-average precision of 0.864 (95% CI: 0.831-0.897), and weighted-average F1-score of 0.829 (95% CI: 0.782-0.876) in the external set, outperforming the performance of the neurologist group.

**Conclusion:** The DL model based on SWS can detect CMBs and classify CSVD, thereby assisting neurologists in optimizing CSVD management.

## Introduction

Cerebral small vessel disease (CSVD) is a chronic and progressive vascular disease affecting capillaries, arterioles, and small veins supplying brain deep structures, often leading to cerebral hemorrhage, dementia, and stroke.^1, 2^ CSVD comprises arteriolosclerosis (aSVD) related to age or vascular risk factors, cerebral amyloid angiopathy (CAA) caused by vascular deposition of β-amyloid, and cerebral autosomal dominant arteriopathy with subcortical infarcts and leukoencephalopathy (CADASIL) caused by *NOTCH3* gene mutations which encode a vascular smooth muscle transmembrane protein involved in vascular development and smooth muscle cell differentiation.^3, 4^ The current treatment strategy for CSVD typically involves secondary stroke prevention,^5, 6^ focusing on prevalent types, such as aSVD and CAA. The role of CADASIL, a typically inherited small vessel disease, in stroke prevention should not be ignored.^7^ However, there is controversy regarding whether the secondary prevention measures for stroke are suitable for these types of CSVD; there is no uniform treatment standard.^3^ For example, antithrombotic therapy is generally not recommended for treating CAA because amyloid angiopathy is an independent risk factor for hemorrhage.^8, 9^ Patients with CADASIL should avoid using antiplatelet drugs to prevent an increased risk of hemorrhage,^10, 11^ antiplatelet therapy should not be withheld for patients with aSVD, and may even be beneficial for the prevention of lacunar stroke.^6, 12^ Optimizing the management of CSVD depends on the individual patient’s intracerebral hemorrhage risk and the type of CSVD. Microbleeds (CMBs), which manifest as small, oval, hypointense lesions on brain susceptibility-weighted MR sequences [SWS, including T2-star-weighted angiography (SWAN) and susceptibility-weighted imaging (SWI)], serve as indicators for evaluating the risk of intracerebral hemorrhage.^13, 14^ CMBs also serve as neuroimaging biomarkers for CSVD and play a critical role in the diagnosis of CSVD, especially CAA.^13, 15-17^ Hence, accurate identification of CMBs is useful for evaluating the risk of intracerebral hemorrhage; facilitating classification of CSVD based on the presence of CMBs, both of which are beneficial for optimizing CSVD management.

Recently, there has been a keen interest in the application of deep learning (DL) to medical images. Several studies indicate that DL can reliably detect lesions and diagnose diseases, such as detecting primary bone tumors on radiographs and diagnosing muscular dystrophies using MRI.^18, 19^ The clinical application of these models may improve the reliability and accuracy of lesion assessment or disease diagnosis, potentially leading to improved diagnostics and better treatment. Therefore, in this study, we used SWS data from patients with three types of CSVD collected from three independent centers to develop and test an end-to-end, two-task DL model that can detect CMBs and classify CSVD.

## Methods

### Patients selection

The Medical Ethics Committee of the Third Affiliated Hospital of Sun Yat-sen University approved this retrospective multicenter study. This study was conducted following the Declaration of Helsinki. Requirements for informed consent were waived because of the study’s retrospective nature.

In this study, 364 CSVD patients (217 patients with aSVD, 106 patients with CAA, and 41 patients with CADASIL) treated at three independent centers, Third Affiliated Hospital, Sun Yat-sen University (SYSUTH), the Maoming People’s Hospital (MMPH), and the First Affiliated Hospital of SHANTOU University Medical College (STUMFH), were enrolled between January 2017 and May 2022 All patients with aSVD had vascular risk factors, such as hypertension, and their MRI neuroimaging met the STandards for ReportIng Vascular changes on nEurouimaging (STRIVE) for CSVD.^3, 13, 20^ Patients with CAA met the diagnostic criteria for probable CAA according to the Boston criteria version 2.0.^15, 16^ CADASIL was confirmed by a genetic diagnosis of *NOTCH3* gene mutation or a granular osmiophilic material identified in a skin biopsy.^21, 22^ Detailed inclusion criteria can be found in the supplementary materials.

### Dataset curation

SWS of enrolled patients were collected to develop and test the model, regardless of SWS parameters, to improve robustness and applicability. The SWS acquisition details are shown in the supplementary materials (**Table S1**). SWS of patients from SYSUTH (a hospital in Guangzhou city) and MMPH (a hospital in Maoming city) served as the development set, and SWS of patients from STUMFH (a hospital in Shantou city) served as the external test set for testing the geographic performance of the model.^23^ Additionally, the 10-fold cross-validation was used to evaluate the stability of the DL model.

### Image of SWS preprocessing

The SWS images were pre-processed before inputting the DL model. Firstly, the orientation of all SWS was uniformly adjusted to left-posterior-superior, and the brain area was extracted from the irrelevant background. Subsequently, the pixel value in the image was ranked, and those topping ≥ 99.5% were denoted as *x*_*u*_. Finally, the pixel value of the entire image was normalized and scaled (*X* = *x*/*x*_*u*_).

### Annotation procedures

To train the model for detecting CMBs, an author (R.Z.W), who was blinded to patients’ clinical data, performed segmentations of the CMBs using the software ITK-SNAP (version 3.8.0). Another author (B.Q) reviewed the results. To evaluate the reproducibility of the manual segmentations, 30 SWS were randomly selected from the development dataset, and an additional segmentation was performed three months after the initial segmentation.

To train and test the model for classifying CSVD into aSVD, CAA, and CADASIL, all SWS underwent additional annotation procedures to confirm the type of CSVD, i.e., the SWS label. A committee of three senior neurologists (B.Q, Z.Q.L, and H.L) annotated labels based on inclusion criteria, with more resource-intensive neurologist annotations being reserved. In cases where of disagreement regarding a label, the committee discussed and reached a consensus. If consensus could not be achieved, the SWS was excluded. In summary, each SWS label received one committee consensus annotation, which was regarded as the gold-standard for model evaluation. In the external test set, four neurologists (L.W, T.T.L, X.H.H, X.L) that were not part of the committee provided individual annotations for labels, and these labels were utilized to compare the model’s performance.

### Network architecture

Figure 1 shows detailed information regarding our architecture. Mask R-CNN was used to detect CMBs slice-wise and to obtain a semantic segmentation mask for CMBs.^24^ Next, a multi-instance learning (MIL) network was applied to classify CSVD sequence-wise.^25^ Semantic segmentation masks of CMBs generated by Mask-RCNN were then spliced with the original corresponding SWS to form a double-channel image as the model’s input for classifying CSVD, inducing the model to concentrate on representations of CSVD on SWS. Resnet50 served as the backbone to extract representations of SWS on each slice layer by layer. Representations of each slice were exhibited as a 1024-dimension vector. Representations of every slice belonging to the same SWS were aggregated into one feature vector using the attention module, among which those representations of slices relating to the final prediction CSVD type were given higher weights, and vice versa. This feature vector was concatenated with the corresponding patient’s age and sex to generate a 1026-dimension vector, serving as the sequence’s representation. Finally, a fully connected layer was used to predict CSVD type according to sequence representation. Gradient-weighted Class Activation Mapping (Grad-CAM) ^26^ was applied to visualize where the MIL network focuses its attention when developing a prediction.

**Figure 1.**
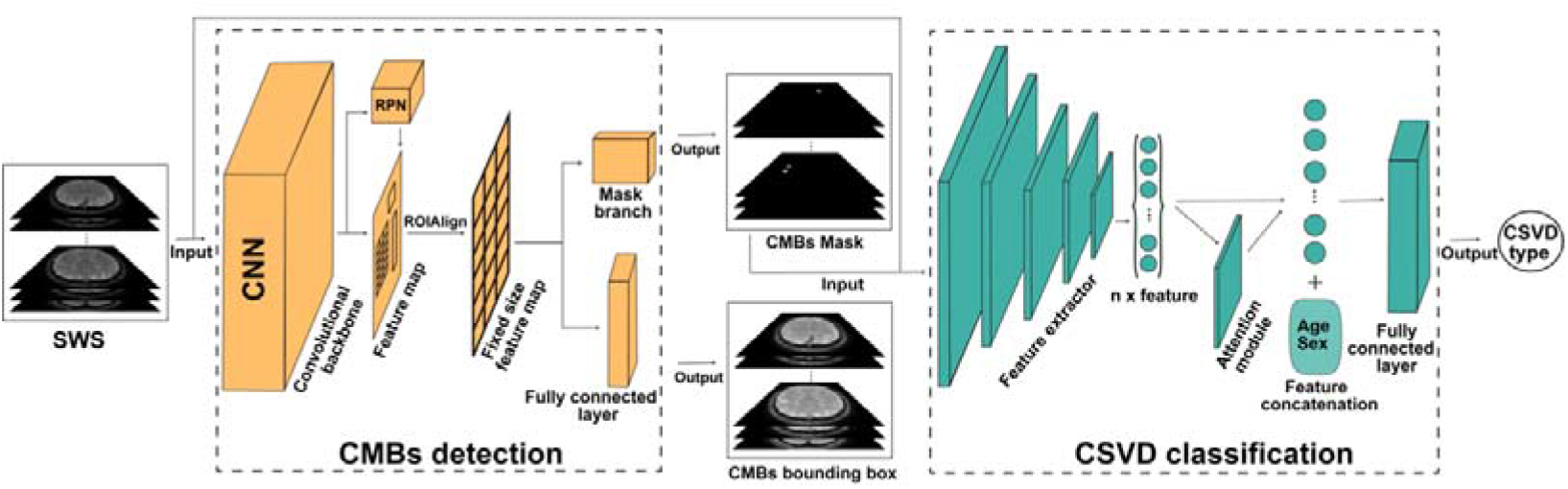
Deep learning architecture overview. Mask R-CNN was used to detect CMBs slice-wise and to obtain a semantic segmentation mask of CMBs. Next, semantic segmentation masks of CMBs were spliced with the originally corresponding SWS to input the MIL network for classifying CSVD SWS-wise. CMBs: microbleeds; MIL: multiple instance learning.

### Model training

Adjacent slices in the SWS were combined to form a three-channel two-dimensional image and input into the Mask-RCNN network to identify CMBs. The manual segmentation of CMBs from the middle channel of the two-dimensional image was used as the ground truth. In CMBs detection training, loss functions used for segmentation and boundary box placement included per-pixel sigmoid loss, binary loss, and regression loss, respectively. During the training for CSVD classification, a balanced random sampling of three categories of CSVD samples was realized using weighted sampling, and the batch size was defined as 1, which guaranteed that only one sample was input into the model. The loss function of the model for CSVD classification was the cross-entropy loss function. The parameters of the model, including the feature extraction module, were trained from end to end using the reverse value of the cross-entropy loss function. The Adam optimizer was used to optimize the parameters of the second model, and the initial learning rate was set at 0.001. An early stopping mechanism was used to control the performance of the training process. When the loss function on the internal test set of the development set did not decline for five consecutive epochs, the training of the model was terminated, and the best model was preserved to avoid a large overfitting of the model on the training set. To classify CSVD, we implemented the 10-fold cross-validation to obtain ten models on the development set. To test the model’s applicability for classifying CSVD, the performance of the ensemble model obtained by aggregating ten models was compared with that of four neurologists (L.W, T.T.L, X.H.H, X.L) on the external test set. The training parameters and source code can be found online (*https://github.com/Huatsing-Lau/CSVD-CMBs-Detection-and-Classification.git*).

An AMD EPYC 7742, 2.25-GHz CPU, and an A100 GPU (Nvidia) were used and run on a Linux system (Ubuntu, version 7.5.0) with a CUDA version 11.7 platform. Model implementation was performed using open-source software (Python, version 3.8.3; Python Software Foundation), Pytorch, version 1.13.0, and torchvision version 0.14.0.

### Statistical analysis

Mean and standard deviation (SD) were used to describe continuous variables, while percentages were used to describe non-continuous variables. Differences between groups were tested using one-way ANOVA and the Chi-square test, respectively. The intersection of union (IoU), Dice score, and recall were calculated to evaluate the model’s performance in detecting CMBs. The IoU was used to assess the performance of bounding boxes. Bounding box placement was considered correct for an IoU above 0.5. The Dice score was used to assess the segmentation performance. Recall was used to evaluate the proportion of the true CMBs predicted by the model. Confusion matrices, receiver operating characteristic curves (ROC), precision, and accuracy were used to evaluate the performance of the model in classifying CSVD. Confusion matrices were used to illustrate the label of CSVD classification, where the model prediction or the individual neurologist predictions were discordant with the committee consensus. ROC analysis was used to calculate the area under the receiver operating characteristic curve (AUC) to assess model discrimination for each label class. Both accuracy, defined as the number of true positive predictions of three categories divided by the sum of samples, and precision, defined as the number of true positive predictions divided by the number of positive predictions were calculated to provide complementary information to the ROC analysis.^27^ The F1 score provides a useful complementary performance measure to the AUC, especially in the context of multi-class prediction, and is less sensitive to class imbalance issues. The weighted-average of the indices was calculated to eliminate the imbalance between the number of categories. Statistical significance was set at P < 0.05. The 95% confidence interval (CI) was calculated for each index. IBM SPSS Statistics (version 26), scikit-learn (version 0.24.2), and the statsmodels (version 0.13.5) were used to analyze the metrics of the models.

## Results

### Patients and datasets characteristics

This study enrolled 364 patients with CSVD, of whom 336 were included in the development set and 28 in the external test set. The development set was comprised of 203 aSVD patients, 99 CAA patients, and 34 CADASIL patients. The mean age (± standard deviation) of the development set patients was 64.88 years ± 6.71 [aSVD (65.34 years ± 6.12); CAA (64.41 years ± 6.98); CADASIL (63.56 years ± 8.93)], and 34.82% (n = 117) of the development set patients were female [aSVD (76/203, 37.44%); CAA (28/99, 28.29%); CADASIL (13/34, 38.24%)]. The external test set consisted of 14 aSVD patients, 7 CAA patients, and 7 CADASIL patients The mean age (± standard deviation) of the external test set patients was 64.18 years ± 6.83 [aSVD (64.36 years ± 6.05); CAA (66.57 years ± 10.16); CADASIL (61.42 years ± 3.41)], and 32.14% (n = 9) of the external test set patients were female [aSVD (3/14, 21.43%); CAA (3/7, 42.86%); CADASIL (3/7, 42.86%)] (Table 1). The SWS in the development set came from 6 different MRI scanners, and the SWS in the external test set came from another MRI scanner. The MRI scanner in the external test set differed from any scanner in the development set. MRI scanners and SWS parameters are summarized in supplementary materials (Table S1).

**Table 1.** Patients and datasets characteristics

| Characteristic | Overall | Development set | External testing set |
| --- | --- | --- | --- |
| number of patients | 364 | 336 | 28 |
| age (y ± SD) * | 64.83 ± 6.72 | 64.88 ± 6.71 | 64.18 ± 6.83 |
| female sex (%) | 126 (34.62%) | 117 (34.82%) | 9 (32.14%) |
| CSVD type |  |  |  |
| aSVD (%) | 217 (59.62%) | 203 (61.42%) | 14 (50.00%) |
| CAA (%) | 106 (29.12%) | 99 (29.46%) | 7 (25.00%) |
| CADASIL (%) | 41 (11.26%) | 34 (10.12%) | 7 (25.00%) |

**Note.** \* mean ± standard. The numbers in parentheses are the percentages. Unless otherwise specified the data indicate the number of participants. The SWS of patients from SYSUTH and MMPH served as the development set, and the SWS of patients from STUMFH served as the external test set for testing the geographic performance of the model. The 10-fold cross-validation was used to evaluate the stability of the DL model.

### CMBs detection

The reproducibility of the manual annotation of CMBs was calculated with an IoU of 0.820 ± 0.039 for bounding box placement, and a Dice score of 0.900 ± 0.024 for segmentation.

On the internal test set of the development dataset, the performance of the model for detecting CMBs presented an IoU of 0.594 ± 0.258 for bounding box placement, and a Dice score of 0.709 ± 0.230 for segmentations. The proportion of the true CMBs predicted by the model had a recall of 0.879 ± 0.228 in the internal test set of the development set. On the external test set, the performance of the model in detecting CMBs presented an IoU of 0.523 ± 0.319 for bounding box placement, and a Dice score of 0.627 ± 0.296 for segmentations. The proportion of the true CMBs predicted by the model had a recall of 0.706 ± 0.365 in the external test set. Pearson analysis to evaluate the correlations of CMBs areas predicted by the model and the ground truth indicated correlations of 0.832 and 0.797 in the internal test set of the development set and external test set, respectively (Figure 2 A, B).

**Figure 2.**
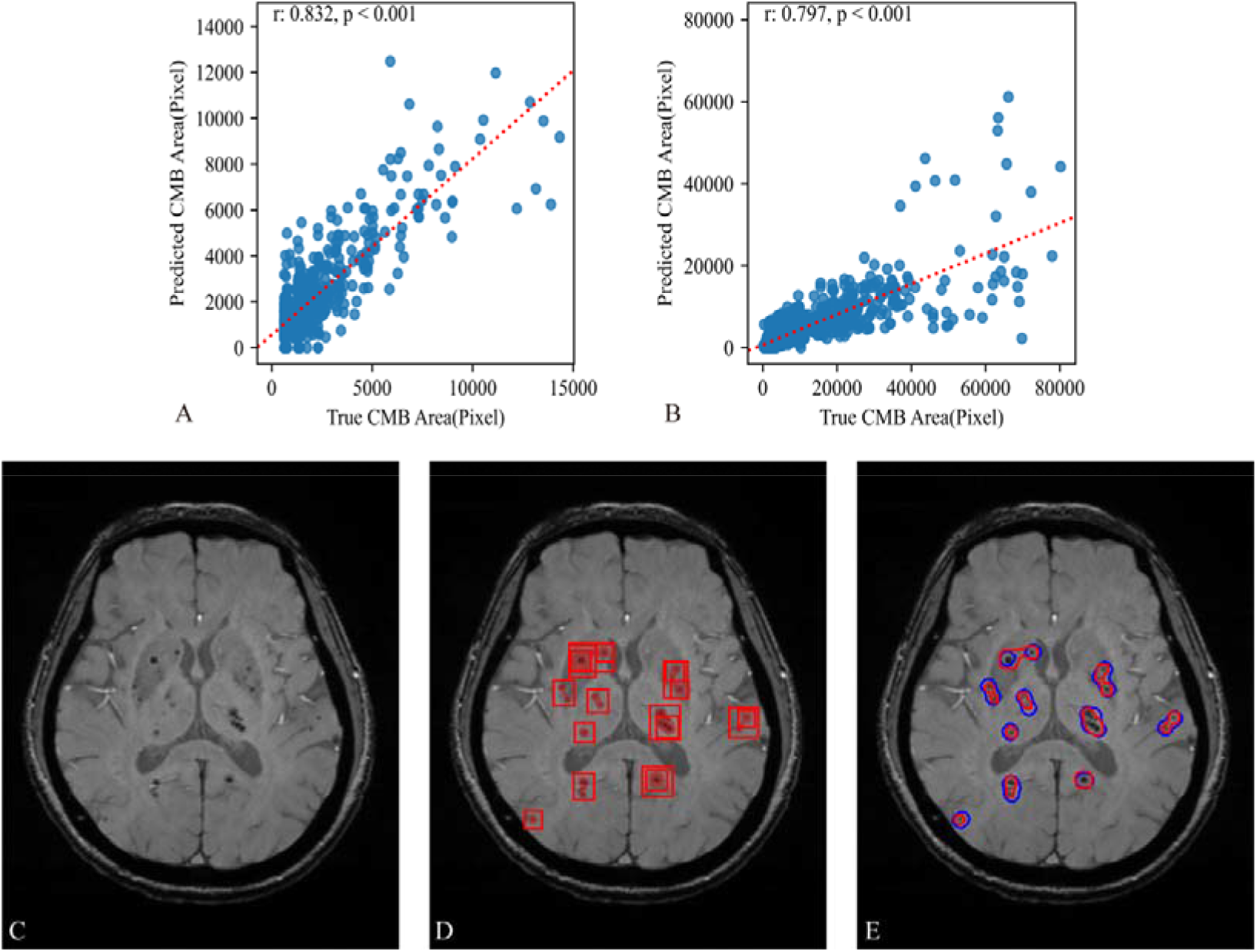
Graphs show scatter diagrams of the Pearson analysis and an example of CMBs detection. (A) The analysis yielded a correlation coefficient of 0.832 for the internal test set of the development set. (B) The analysis yielded a correlation coefficient of 0.797 for the external test set. (C) Original SWS image. (D) Bounding box placement and segmentation of CMBs performed by the Mask R-CNN. (E) Comparison between the CMBs region identified by the model (red) and the groundtruth (blue).

### CSVD classification

On the internal test set of the development set, the performance of the model for classifying CSVD into aSVD, CAA, and CADASIL across 10-fold cross-validation presented a mean weighted-average AUC of 0.865 (95% CI: 0.841-0.884), a mean accuracy of 0.732 (95% CI: 0.718-0.746), a mean weighted-average precision of 0.755 (95% CI: 0.743-0.768), and a mean weighted-average F1-score of 0.733 (95% CI: 0.719-0.747) (Table 2). ROC analysis showed a mean AUC for categorizing CSVD into aSVD, CAA, and CADASIL of 0.867 (95% CI: 0.805-0.929), 0.863 (95% CI: 0.802-0.924), and 0.834 (95% CI: 0.773-0.895), respectively, in the internal test set of the development set (Figure 3 A). On the external test set, the performance of the model at classifying CSVD into aSVD, CAA, and CADASIL across 10-fold cross-validation presented a mean weighted-average AUC of 0.899 (95% CI: 0.884-0.914), a mean accuracy of 0.717 (95% CI: 0.704-0.731), a mean weighted-average precision of 0.717 (95% CI: 0.702-0.733), and a mean weighted-average F1-score of 0.705 (95% CI: 0.691-0.719) (Table 2). ROC analysis showed a mean AUC for categorizing CSVD to aSVD, CAA, and CADASIL of 0.954 (95% CI: 0.946-0.970), 0.929 (95% CI: 0.916-0.940), and 0.760 (95% CI: 0.734-0.788), respectively, in the external test set (Figure 3 B). The results for each run of 10-fold cross-validation of the model for CSVD classification are summarized in supplementary materials (Table S2).

**Table 2.** Metrics in CSVD classification for the model and the neurologist group

| Metrics | Internal test set | External test set |  |  |
| --- | --- | --- | --- | --- |
|  | mean of 10-told cross-validation | mean of 10-told cross-validation | ensemble model | neurologist group |
| weighted-average AUC | 0.865<br>(0.841-0.884) | 0.899<br>(0.884-0.914) | 0.908<br>(0.895-0.921) | \ |
| accuracy | 0.732<br>(0.718-0.746) | 0.717<br>(0.704-0.731) | 0.819<br>(0.768-0.870) | 0.759<br>(0.740-0.779) |
| weighted-average precision | 0.755<br>(0.743-0.768) | 0.717<br>(0.702-0.733) | 0.864<br>(0.831-0.897) | 0.767<br>(0.747-0.786) |
| weighted-average F1-score | 0.733<br>(0.719-0.747) | 0.705<br>(0.691-0.719) | 0.829<br>(0.782-0.876) | 0.762<br>(0.742-0.781) |

**Note.** the numbers in parentheses are the 95% confidence interval.

**Figure 3.**
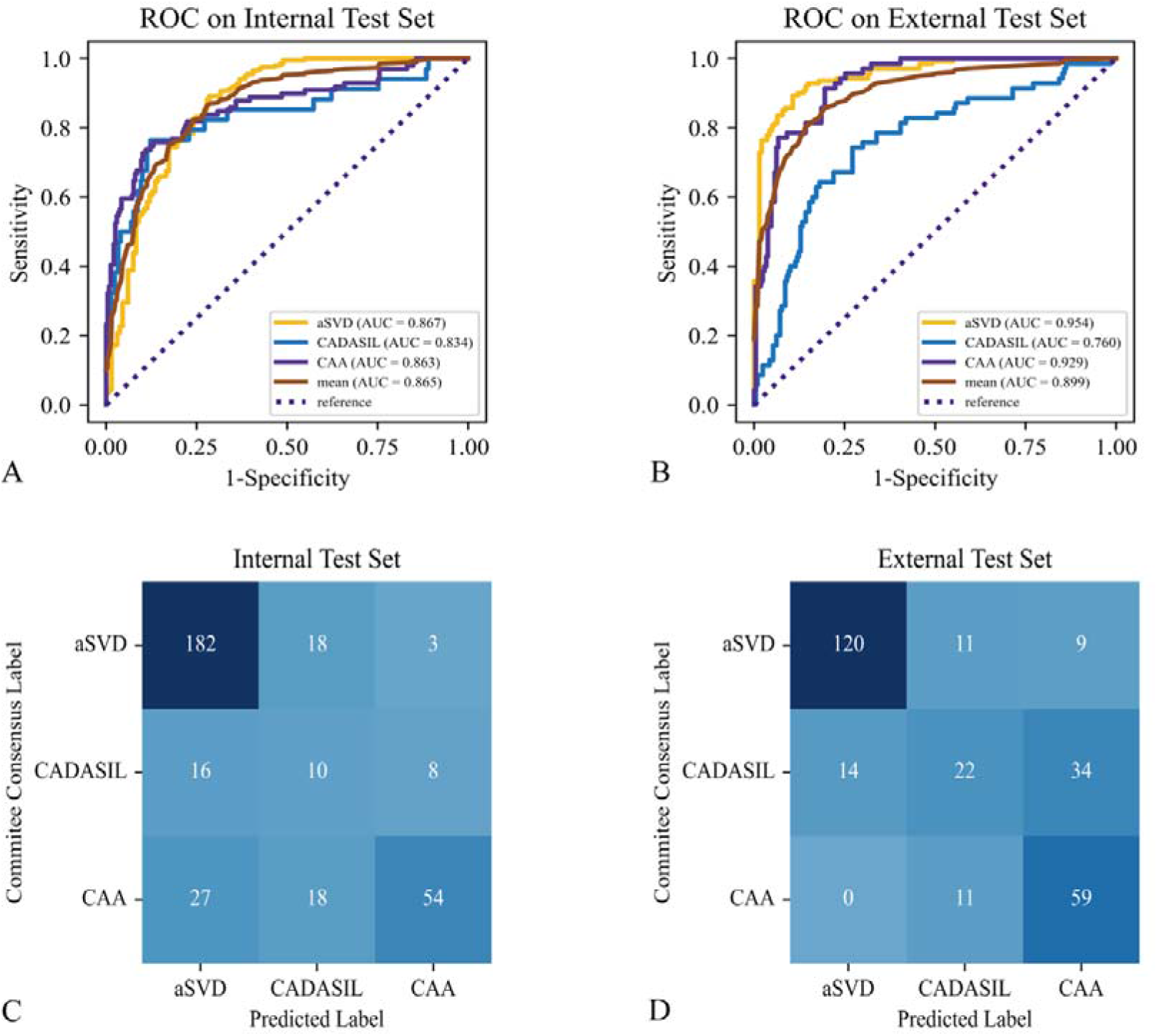
Graphs show the ROC and confusion matrices for evaluating the model performance in 10-fold cross-validation in classifying CSVD. (A-B) ROC for model performance in 10-fold cross-validation on the internal test set of the development set and the external test set, respectively. (C-D) Confusion matrices for model performance in 10-fold cross-validation on the internal test set of the development set and the external test set, respectively. Mean represents the weighted-average AUC.

In the external test set, the performance of the ensemble model for classifying CSVD presented a weighted-average AUC of 0.908 (95% CI: 0.895-0.921), an accuracy of 0.819 (95% CI: 0.768-0.870), a weighted-average precision of 0.864 (95% CI: 0.831-0.897), and a weighted-average F1-score of 0.829 (95% CI: 0.782-0.876) (Table 2). ROC analysis showed an AUC for categorizing CSVD to aSVD, CAA, and CADASIL of 0.959 (95% CI: 0.948-0.969), 0.946 (95% CI: 0.932-0.959), and 0.769 (95% CI: 0.731-0.805), respectively, in the external test set (Figure 4). Comparatively, the performance of the group of neurologists in classifying CSVD into aSVD, CAA, and CADASIL on the external test set was inferior to that of the ensemble model, with an accuracy of 0.759 (95% CI: 0.740-0.779), weighted-average precision of 0.767 (95% CI: 0.747-0.786), and weighted-average F1-score of 0.762 (95% CI: 0.742-0.781). The performance of each neurologist for CSVD classification on the external test set is summarized in supplementary materials (Table S3 and Figure S1). Figure S2 shows an example of an aSVD SWS superimposed on a Grad-CAM activation map.

**Figure 4.**
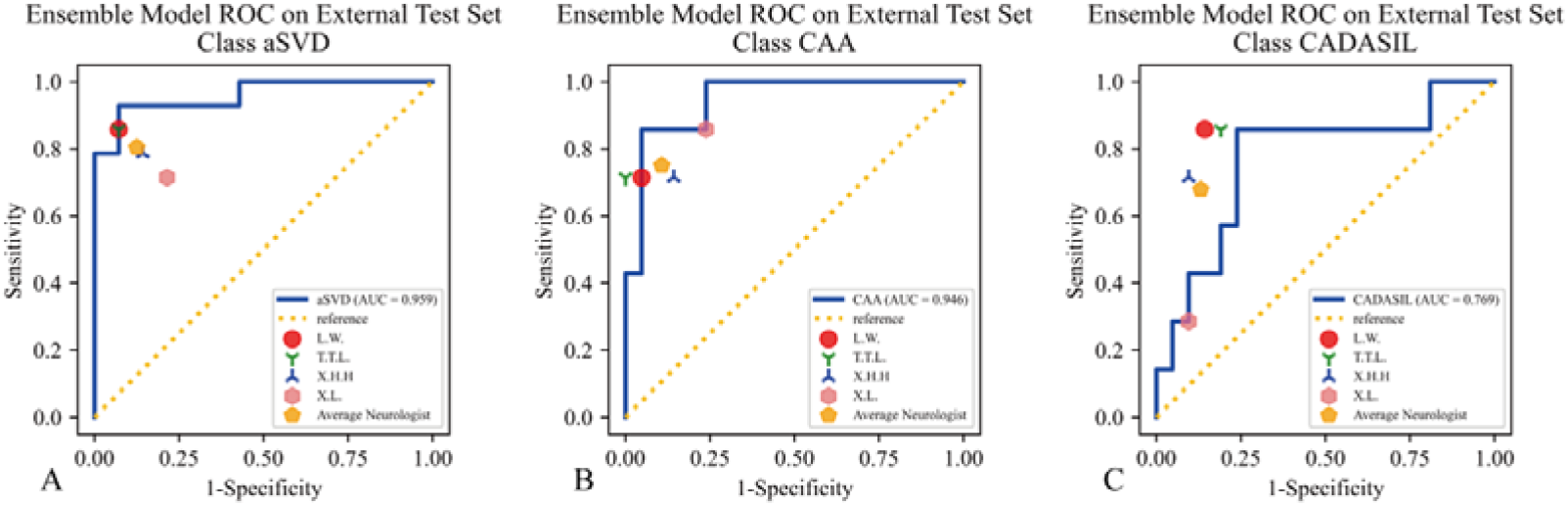
ROC for evaluating the ensemble model performance in classifying CSVD on the external test set. Dots of different shapes represent individual neurologist performance and average neurologist performance.

## Discussion

Optimizing CSVD management usually involves assessing the risk of intracerebral hemorrhage and determining the type. However, accurately evaluating these factors using MRI, which is the primary tool for examining and diagnosing CSVD, can be inconvenient. Firstly, it is difficult to visualize CMBs, indicators for evaluating the risk of intracerebral hemorrhage,^14^ on SWS because of their small size and the extensive co-occurrence of similar objects in the brain, such as blood vessels and calcifications, which can be misinterpreted as CMBs. Hence, the visual inspection of CMBs on SWS requires significant expertise and can be time-consuming and prone to errors. Secondly, differentiating CSVD based on MRI results can be challenging since the diagnosis of CSVD primarily relies on neuroimaging biomarkers and clinical presentation, which may not have clear-cut categorical distinctions.^3^ In addition, comprehensive diagnosis of CSVD typically requires multiple MR sequences,^13^ and further biopsy or genetic testing is necessary for confirmation, such as in cases of CAA and CADASIL.^10, 15^ Both can not only increase the clinical workload but also impose a financial burden on patients. Herein, we used SWS to develop and test a DL model that can detect CMBs and classify CSVD into aSVD, CAA, and CADASIL to assist neurologists in optimizing the management of CSVD.

Some recent studies have used DL to detect CMBs on SWS,^28-30^ but images were cropped manually around the CMBs, or data were collected from only one center, thus, possibly limiting the clinical applicability and reliability. Moreover, previously proposed models have adopted non-end-to-end algorithms for detecting CMBs, which could potentially result in low efficiency and human errors. In this study, we proposed to use Mask R-CNN, an end-to-end algorithm, to detect CMBs slice-wise, which was developed and tested using SWS collected from three independent centers, regardless of the MRI system platforms or parameters. Using heterogeneous data to develop and test the model indicates that our model has stronger applicability and robustness, and using an end-to-end algorithm for CMBs detection indicates that our model has higher efficiency and fewer human errors. Additionally, another significance of detecting CMBs in this work is the use of the CMBs segmentation masks generated by the model to assist with the second task, i.e., CSVD classification. Performing two tasks to capture more detailed information can improve the model stability.^31^

There are several studies on the application of DL in CSVD ^32-34^, but the focus is on the biomarkers rather than aetiopathogenic classification, limiting the benefit of optimizing CSVD management. Herein, we proposed an SWS-based MIL network that can classify sequence-wise CSVD into aSVD, CAA, and CADASIL from the perspective of etiopathogenesis. The MIL network must be able to extract SWS representations relative to the above types of CSVD to accomplish this task. SWS is presented in a three-dimensional form with a large volume, making it reasonable to use a three-dimensional convolutional neural network (3D-CNN) to extract representations. However, using a 3D-CNN may be time-consuming and costly, leading to low calculation efficiency. Reslicing can be used to decrease the resolution of the SWS, thereby improving its efficiency. However, high resolution is required to recognize CSVD image biomarkers such as CMBs; therefore, we abandoned the method of lessening SWS volume by reslicing. Here, we used a two-dimensional convolutional neural network (2D-CNN) to extract SWS representations instead of 3D-CNN. The main limitation of 2D-CNN is that it is applicable only to two-dimensional but not three-dimensional images composed of multiple ordered slices, such as SWS.^35^ To overcome this limitation, we used the attention module to aggregate representations of each SWS slice into the whole SWS representation, inspired by the interpretable weakly supervised DL method for data-efficient whole-slide image processing and learning proposed by *Ming et al* ^36^. Besides, to effectively train the model to classify CSVD based on SWS, we used CMBs segmentation masks generated by the Mask R-CNN to induce the MIL network to concentrate on the representations of the above types of CSVD. In 10-fold cross-validation, the model demonstrated consistent performance on both internal and external test set, indicating that our model has excellent generalizability. Based on the Grad-CAM activation map, regions within the brain parenchyma were found to have a high degree of contribution to the model’s ability to develop CSVD prediction, indicating that our model relies on brain representations to make differential decisions, further supporting its stability. Additionally, the model achieved better performance than the neurologist group in classifying CSVD into aSVD, CAA, and CADASIL on the external test set, indicating its potential to determine CSVD types using only SWS and providing an effective approach for clinical differential diagnosis of CSVD, especially for generalized medical institutions lacking adequate diagnostic equipment. Furthermore, the model also offers the added advantage of confirming CAA or CADASIL patients without the need for biopsy or genetic testing, thereby decreasing the financial burden on patients.

This study has some limitations and potential areas for future research. First, given the rarity of monogenic CSVD,^10, 37^ the present study included a relatively small number of patients with CADASIL and did not include other types of CSVD, such as inflammatory vasculitides. To overcome this limitation, future studies should aim to recruit more diverse and extensive cohorts. Second, because the SWS labels used to train and test the model for classifying CSVD are annotated by a committee based on neuroimaging biomarkers and clinical symptoms, there may be random human errors, especially in annotating aSVD and CAA; thus, the performance of our model in classifying CSVD is based on the neurologist-level “gold-standard”, rather than a true gold-standard. In future research, molecular and protein-level studies need to be combined to eliminate human errors in the model’s performance. Third, we did not include cases with mixed pathologies, such as small vessel disease combined with inflammatory vasculitides, because we considered that the use of neuroimaging biomarkers and symptoms was insufficient to distinguish these mixed etiologies. In future research, more clinical information, such as biochemical examinations, should be included to train and test the model and achieve better results.

Overall, we developed and tested a DL model based on SWS that can detect CMBs and presented an IoU of 0.523±0.319, a Dice score of 0.627±0.296, and a recall of 0.706±0.365, classification of CVSD into aSVD, CAA, and CADASIL had a weighted-average AUC of 0.908 (95% CI: 0.895-0.921), an accuracy of 0.819 (95% CI: 0.768-0.870), a weighted-average precision of 0.864 (95% CI: 0.831-0.897), and a weighted-average F1-score of 0.829 (95% CI: 0.782-0.876), providing a promising approach to assist neurologists in optimizing the management of CSVD.

## Data Availability

The Medical Ethics Committee of the Third Affiliated Hospital of Sun Yat-sen University approved this retrospective multicenter study.

https://github.com/Huatsing-Lau/CSVD-CMBs-Detection-and-Classification.git

## Non-standard Abbreviations and Acronyms

CMBs: microbleeds
CSVD: cerebral small vessel disease
DL: deep learning
SWS: susceptibility-weighted MR sequence
aSVD: arteriolosclerosis
CAA: cerebral amyloid angiopathy
CADASIL: cerebral autosomal dominant arteriopathy with subcortical infarcts and leukoencephalopathy
MIL: multi-instance learning
IoU: intersection over union
ROC: receiver operating characteristic curve
AUC: area under receiver operating characteristic curve
SWAN: T2-star-weighted angiography
SWI: susceptibility-weighted imaging
STRIVE: STandards for ReportIng Vascular changes on nEurouimaging
SYSUTH: Third Affiliated Hospital, Sun Yat-sen University (SYSUTH)
MMPH: Maoming People’s Hospital
STUMFH: First Affiliated Hospital of SHANTOU University Medical College
SD: standard deviation
Grad-CAM: Gradient-weighted Class Activation Mapping
CI: confidence interval
3D-CNN: three-dimensional convolutional neural network
2D-CNN: two-dimensional convolutional neural network

## Acknowledgments

The authors thank *Tingting Lu* for comparing with the model in terms of CSVD classification on the external test set.

## Supplemental material

Supplemental methods

Table S1-S3

Figure S1-S2

